# COVIDTrach; a prospective cohort study of mechanically ventilated COVID-19 patients undergoing tracheostomy in the UK

**DOI:** 10.1101/2020.10.20.20216085

**Authors:** COVIDTrach collaborative, NJI Hamilton, AGM Schilder, T Jacob, G Ambler, M Singer, MM George, F Green, R Vasanthan, J Goulder, E Jackson, A Arora, N Kumar, C Schilling, S Laha, I Ahmad, B McGrath, MA Birchall, NS Tolley, G Sandhu, T Tatla, N Sharma, P Stimpson, P Andrews, N Mercer, P Nankivell, O Breik, P Praveen, M Idle, T Martin, S Parmar, P Pracy, C Jennings, J Higginson, K Fan, E Yeung, J Osher, R Bentley, C Huppa, P Stenhouse, K Hussain, S Hodges, F Ryba, P Surda, EK Bhargava, N Amin, J Collins, M Kelly, D Ranford, A Takhar, C Tornari, M Verkerk, C Xie, D Pennell, C Al-Yaghchi, L Ritchie, M Jaafar, M Rouhani, M Ashcroft, N Cereceda-Monteoliva, A Holroyd, J Ng, R Mistry, K Ghufoor, E Warner, H O’Mahony, S Shepherd, N Bhatti, H Drewery, J Hadley, A Mulcahy, H Wilson, R Bhandari, M Griffiths, T Magos, I Balasundaram, M Heliotis, A Loizidou, D York, R Exley, KA Solanki, P Shah, P Kirticumar, A Shah, S Shannon, A Shirazian, Y Bhatt, K Dhadwal, GM Jama, Z Abdi, T Exall, I Ekpemi, R Roplekar-Bance, C Walker, N Glibbery, K Karamali, A Li, A Rovira, D Dawson, T Munroe-Gray, P Sethukumar, J Phillips, A Williamson, R Saha, M Roberts, H Lee-Six, B Misztal, S Millington, M Musalia, S Suresh, A Cardozo, M Dunbobbin, A Tse, S Shahidi, M Chachlani, K Jolly, J Fussey, M Misurati, M Osborne, S Ashok, H Aboulgheit, S Khwaja, R Anmolsingh, C Smyth, B Al-Dulaimy, E Omakobia, J Collier, T Browning, A Courtney, P Ward, L Lignos, C Lockie, P Twose, J Heyman, S Berry, P Bishop, D Kathwadia, T Hwara, A Williamson, A Kumar, O Judd, W Parker, TP Davis, T Stubington, T Ali, A Schache, H Koumoullis, E Willcocks, L Skeely, G Dempsey, K Liatsikos, B Borgatta, J Rodrigues, A Glossop, J Sen, N Lawrence, S Bennett, L Wren, V Politidis, D Dhariwal, S Winter, A Kara, T Hunt, G Tattersall, W Udall, B Hill, S Saha, L Bates, C Smart, D Park, R O’Brien, L Linhartova, P Kirkland, J Staufenberg, K Valchanov, H Buglass, U Sheikh, E Tam, J Williamson, A McGrath, S Siddiq, NW Wahid, H Griffiths, M De, A Amlani, P Deutsch, K Markham, C Hall, S Webster, O Barker, P Sykes, A Gupta, A Easthope, S Glaze, B Morris, D Bondin, D Thorley, K Kapoor, S Sirajuddin, S Fang, F van Damme, O Mattoo, E Paramasivam, E Kershaw, S Dewhurst, S Blakeley, C Chivers, L Lindsey, DJ Lin, A Burns, A Wilson, N Macartney, F Franco, K Goodwin, B Cosway, R Glore, H Cunniffe, M Keil, S Burrows, D Moult, D Zolger, J Bakmanidis, D Nair, S Kandiah, M Anwar, A Pericleous, C Hogan, R Temple, D Whitmore, R Sheikh, R Pinto, C Cook, J Broad, U Nagalotimath, E El-Tabal, S Ghaffar, M Dallison, E Leakey, R Harris, J Blair, E France, O Sanders, P Mukherjee, A Gomati, L Moir, CB Groba, C Davies-Husband, N Seymour, S Mahalingam, D Williams, R Lovett, J Lunn, A Armson, A Balfour, K Steele, K Hilliard, S Ladan, P Paul, P Tsirevelou, V Ratnam, H Turner, N Jain, A Muddaiah, M Celinski, J Smith, J Westwood, J Coakes, R Borg, J McEwan, A Tsagkovits, O Mulla, N Stobbs, G Warner, D Pratap, Z Ghani, J Rocke, S Snape, S Ghosh, A Hassaan, M Cameron, A Daudia, S Menon, S Beckett, R Siau, A Howard, C Lamont, C Blore, C Pearce, D Zakai, S Biswas, R Moorthy, J Bates, P Gill, E Riley, P Bothma, S Meghji, W Rutherford, A Lloyd, A Syndercombe, P Smith, N Keates, V Srinivasan, M Junaid, M Kumar, T Antonio, A Vijendren, V Venkatachalam, I Gonzalez, M Lechner, D Chandrasekharan, A Arya, R Brown, H Jones, D Kumar, R Sykes, B Tehan, A Walker, J Whiteside, F Cooper, A Coombs, G Wong, D Walker, S Dennis, A Hormis, A Eldahshan, L Leach, H Paw, M Colomo-Gonzalez, D Chakravarty, S Sanyal, N Mani, B Ranganathan, H Saeed, S Linton, A Thompson, J Whittaker, N Amiruddin, A Sladkowsk, R Gohil, AK Abou-Foul, J Ahmed, S Kishwan, G Walton, P Naredla, A Al-Ajami, S Wilkinson, S Okhovat, A Menon, S Mustafa, E Carey, N Vallabh, T Davies, A. Alatsatianos, R Townsley

## Abstract

**Purpose:** COVIDTrach is a UK multi-centre prospective cohort study project evaluating the outcomes of tracheostomy in patients with COVID-19 receiving mechanical ventilation. It also examines the incidence of SARS-CoV-2 infection among healthcare workers involved in the procedure.

**Method:** An invitation to participate was sent to all UK NHS departments involved in tracheostomy in COVID-19 patients. Data was entered prospectively and clinical outcomes updated via an online database (REDCap). Clinical variables were compared with outcomes using multivariable regression analysis, with logistic regression used to develop a prediction model for mortality. Participants recorded whether any operators tested positive for SARS-CoV-2 within two weeks of the procedure.

**Results:** The cohort comprised 1605 tracheostomy cases from 126 UK hospitals. The median time from intubation to tracheostomy was 15 days (IQR 11, 21). 285 (18%) patients died following the procedure. 1229 (93%) of the survivors had been successfully weaned from mechanical ventilation at censoring and 1049 (81%) had been discharged from hospital. Age, inspired oxygen concentration, PEEP setting, pyrexia, number of days of ventilation before tracheostomy, C-reactive protein and the use of anticoagulation and inotropic support independently predicted mortality. Six reports were received of operators testing positive for SARS-CoV-2 within two weeks of the procedure.

**Conclusions:** Tracheostomy appears to be safe in mechanically ventilated patients with COVID-19 and to operators performing the procedure and we identified clinical indicators that are predictive of mortality.

**Funding:** The COVIDTrach project is supported by the Wellcome Trust UCL COVID-19 Rapid Response Award and the National Institute for Health Research.

**Trial registration:** The study is registered with ClinicalTrials.Gov (NCT04572438).

## Introduction

As of 1^st^ September 2020, there were over 33 million cases of COVID-19 globally with average daily new cases exceeding 250,000 [1]. Data indicate that 5-12% develop a severe illness requiring critical care, of whom 72-81% require invasive mechanical ventilation [2-6]. The UK experienced a peak in infection during the months of March and April 2020 followed by a decline and low plateau of new cases during June and July. Of approximately 10,894 patients recorded in the UK Intensive Care National Audit & Research Centre (ICNARC) database that were admitted to intensive care during this period, over 7792 required advanced respiratory support [7].

Standard UK intensive care practice is to consider tracheostomy after 7-10 days of invasive mechanical ventilation to aid weaning, facilitate comfort and minimize complications relating to the prolonged presence of an oral endotracheal tube [8-11]. The role of tracheostomy in COVID-19 remains controversial. Guidance early in the pandemic ranged from avoiding tracheostomy completely to delaying tracheostomy until 14-21 days after intubation [12-14], and to only proceed once the patient was COVID-19 test negative [15-17]. These recommendations aimed to prevent nosocomial infection among healthcare professionals and to avoid futile procedures. The evidence base for these recommendations was largely based on expert opinion. Subsequent evidence suggests the risk of transmission declines 3-4 days after symptom onset and that standard COVID-19 testing does not reflect the risk of infectivity later in the disease process [18, 19]. Several series reporting outcomes following tracheostomy in ventilated COVID-19 patients demonstrate high rates of survival and weaning success and no SARS-COV-2 infection among those involved in the procedure [20-22].

COVIDTrach is a UK multidisciplinary collaborative project examining the outcomes of tracheostomy in mechanically ventilated patients with COVID-19 in UK intensive care units and the occurrence of SARS-CoV-2 infection among operators. An interim report documenting the first 548 patients during the first two months of the pandemic reported a hospital mortality of 12%, weaning success in 52%, and no instances of SARS-CoV-2 infection among operators at the time of writing [23]. This article presents a larger cohort of patients, their long-term outcomes and a further analysis of parameters that may predict survival.

## Methods

### Study design

COVIDTrach is a multicentre prospective cohort study of mechanically ventilated patients with COVID-19 who undergo tracheostomy. An invitation for clinicians to participate in the COVIDTrach project was disseminated via the UK Federation of Surgical Specialty Associations, its various member organizations and the Intensive Care Society to reach all UK departments involved in tracheostomy in COVID-19 patients. Inclusion into the study was also advertised on societal websites and social media. 137 hospital sites agreed to participate across all four nations of the United Kingdom.

### Patient population

Each participating hospital included all consecutive adult patients over 18 years of age with COVID-19 who underwent elective tracheostomy while receiving invasive mechanical ventilation. Infection was identified by a positive viral RNA test on quantitative RT-PCR testing or when strongly suspected on history, laboratory, and radiological findings in the absence of viral RNA test availability or a positive result. Children under the age of 18 years and those undergoing emergency tracheostomy were excluded.

### Clinical indicators and outcome measures

Demographic data, clinical characteristics, SARS-CoV-2 status, ventilatory requirements before tracheostomy, details of the tracheostomy and the use of personal protective equipment (PPE) were all recorded (data dictionary in appendix). Clinical outcomes included complications, mortality, time from tracheostomy to weaning from mechanical ventilation, success, and time to tracheostomy decannulation, and time from tracheostomy to hospital discharge. Successful weaning was defined as being free from pressure support for greater than 24 hours. Participants were also asked to report whether any of the operators performing the procedure tested positive for SARS-CoV-2 within two weeks of the procedure.

### Procedures

Data was collected using an online survey tool (REDCap) with return codes issued to allow participants to update clinical outcomes prospectively. All participants were asked to update the clinical outcomes of their cases by July 1^st^ 2020 and confirm the data were complete. Hospital sites with missing data or data that had not been updated to within the last two weeks of June 2020, were followed up over the course of July and early August. Hospitals with datasets not updated by 15^th^ August 2020 were removed from the study (n=11).

### Ethics and data governance

The study was approved by the Health Research Authority (20/HRA/3766), the Scottish Public Benefit and Privacy Panel and Northern Island’s Privacy Advisory Committee and was registered as a service evaluation or research study at each participating site. All data collected were anonymized and non-identifiable and did not alter the patient’s clinical care. The study is registered with ClinicalTrials.Gov (NCT04572438).

### Statistical analysis

Variables are presented using either mean (SD), median (IQR) or number (percentage), as appropriate. Groups of variables were compared using either t-tests or chi-squared tests, as appropriate. For multivariable regression analysis, all numerical variables, except age, were log-transformed. Logistic regression was used to develop a prediction model for mortality, after imputing missing values using multiple imputation via chained equations. Backwards elimination at the 10% significance level was used to remove unimportant variables from the model. This model was internally validated using 10-fold cross-validation. Model performance (calibration and discrimination) was quantified using the calibration slope and calibration in the large, and ROC curves.

## Results

### Participants

Between 6^th^ April and 26^th^ August 2020, data was received on 1605 tracheostomies from 126 UK hospitals led by a combination of ENT, maxillofacial and intensive care specialists (Figure 1). The number of tracheostomy cases entered by each hospital ranged from 1-106 (mean 12). Across all cases, over 90% of all data points were completed in all but three variables where completeness was 85% or greater (Supplementary table 1). The average patient age was 58 ± 11 years with a 70:30 male to female ratio. More detailed patient demographics and medical history are presented in Table 1 and COVID-19 test status in Table 2.

**Table 1.**
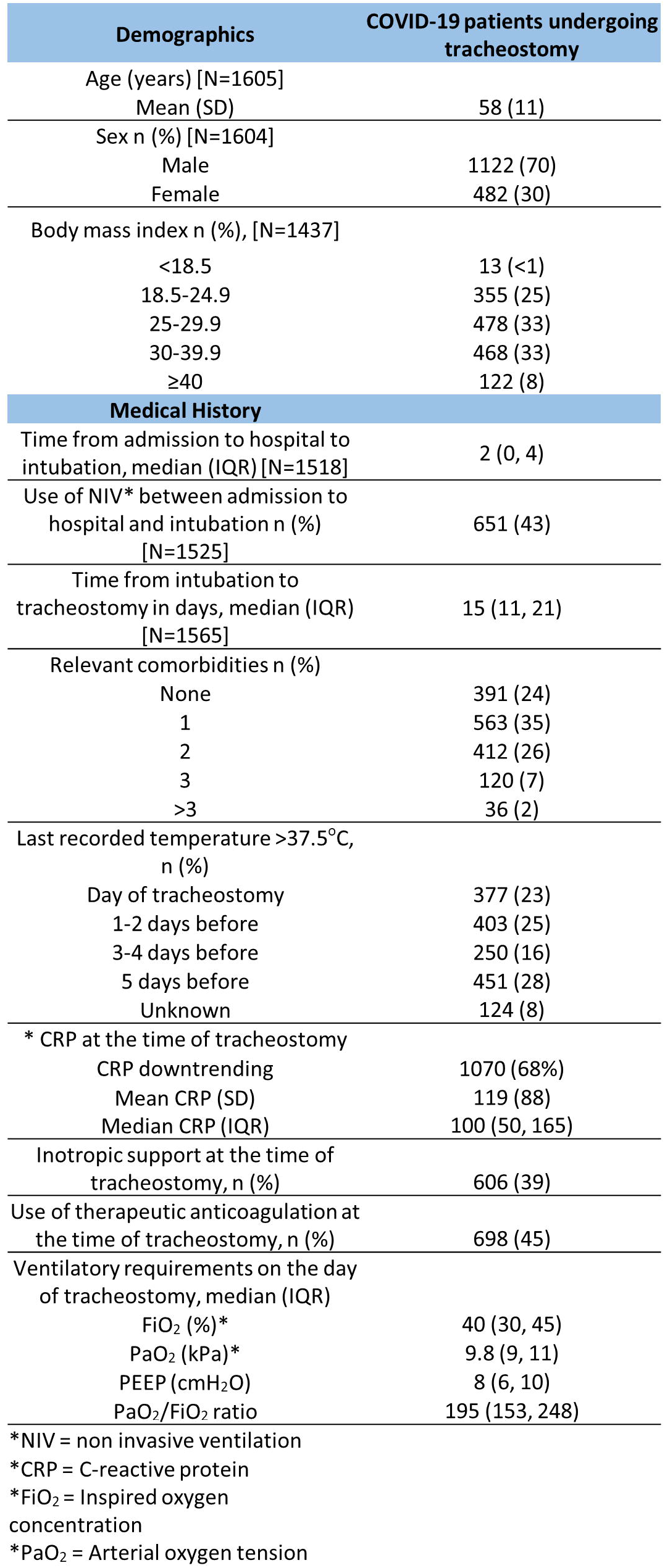
Patient demographics and medical history of 1605 mechanically ventilated patients with COVID-19 who underwent tracheostomy.

**Table 2.**
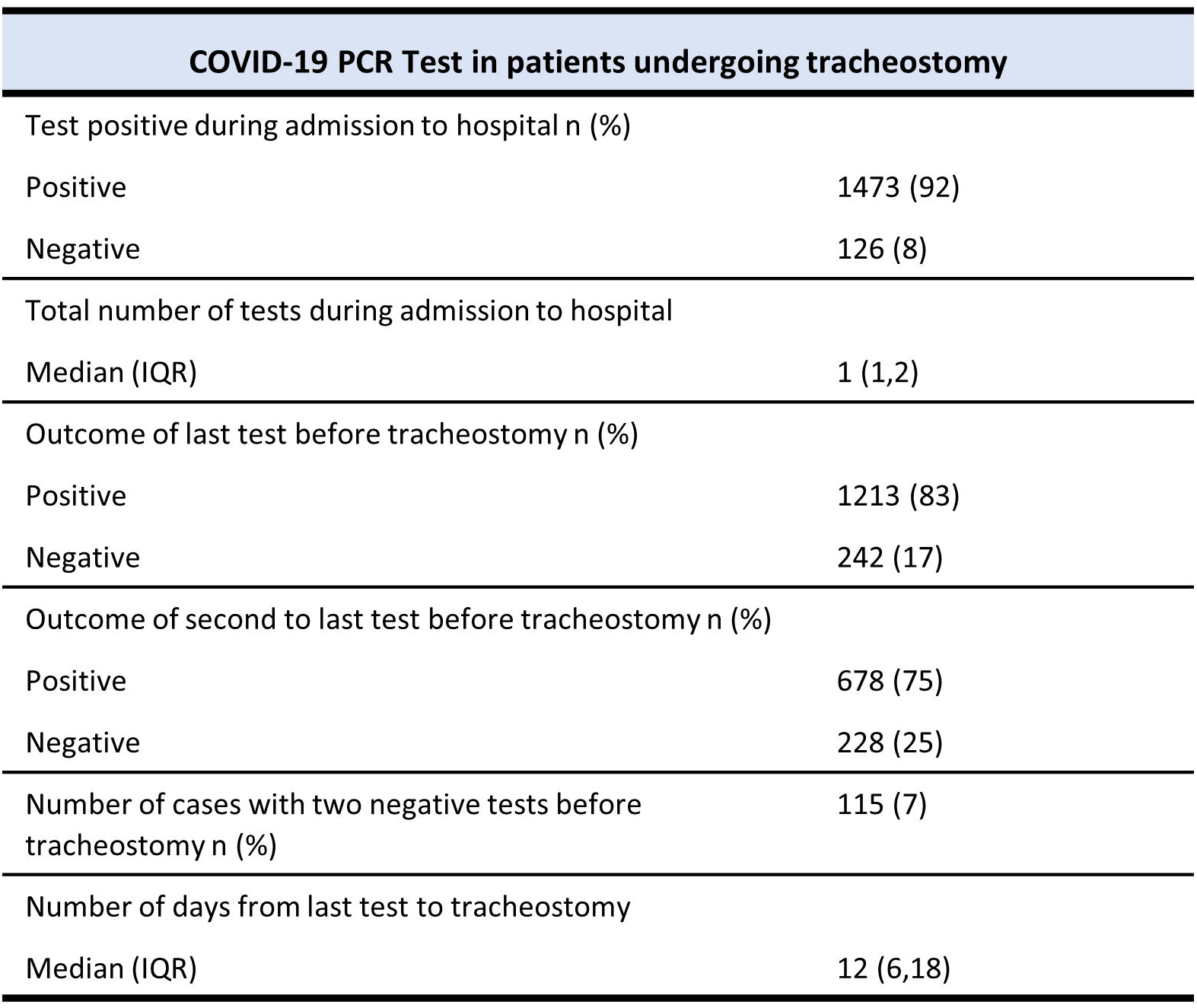
The outcome of COVID-19 PCR testing in 1605 mechanically ventilated patients with COVID-19 undergoing tracheostomy.

**Figure la.**
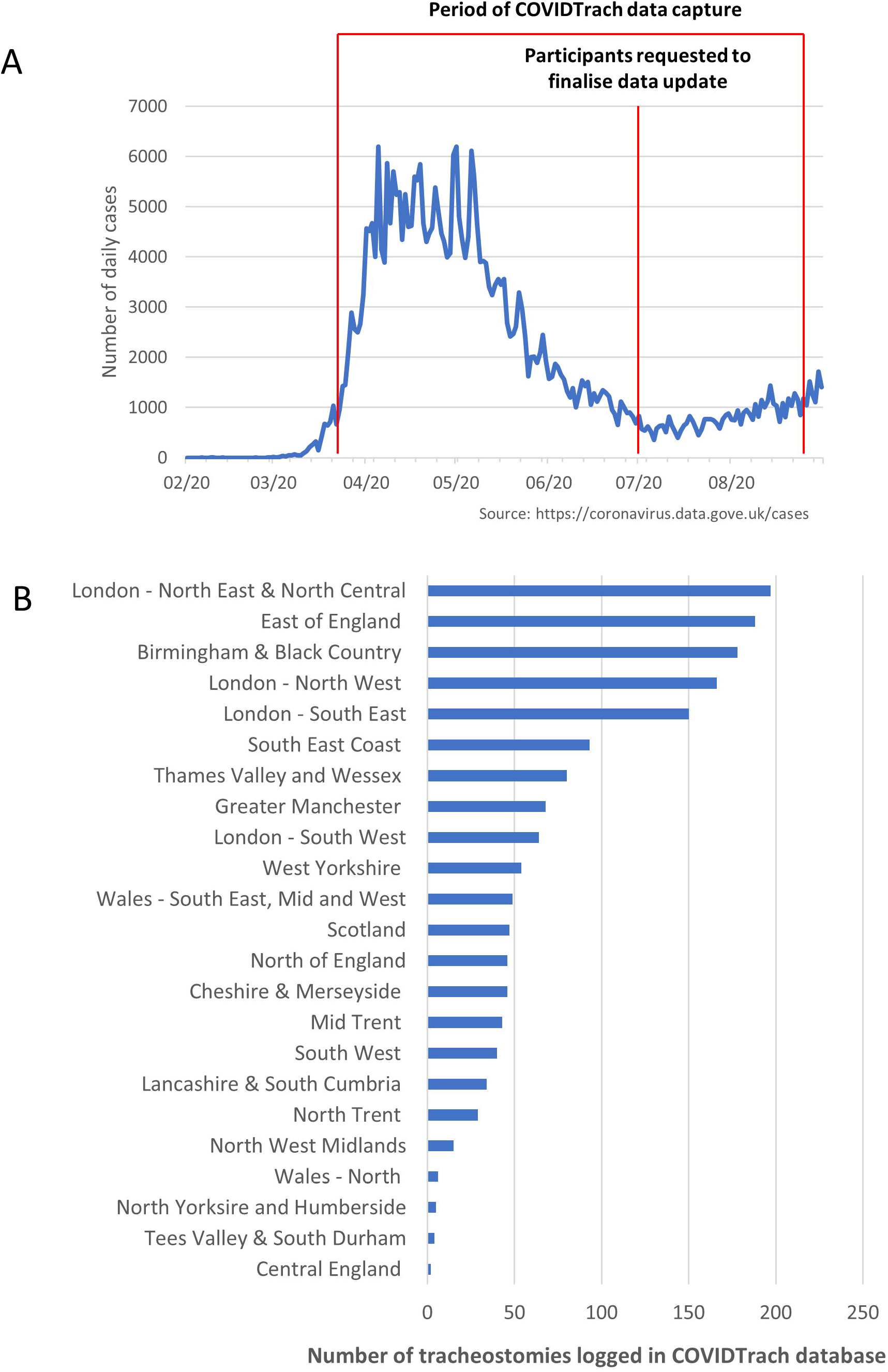
Number of daily cases in the UK from the start of the pandemic to the point of analysis. X-axis represent the month and year. **Figure 1b** Number of tracheostomies logged in the COVIDTrach database by UK region.

### Timing of tracheostomy procedure

The median time from intubation to tracheostomy was 15 days (IQR 11, 21) (Figure 2a). Seventy-three (4.5%) patients had tracheostomy within 4 days of intubation and 227 (14%) after 25 days. The mean C-reactive protein on the day of tracheostomy was 119 (SD, 88) mg/l.

**Figure 2a.**
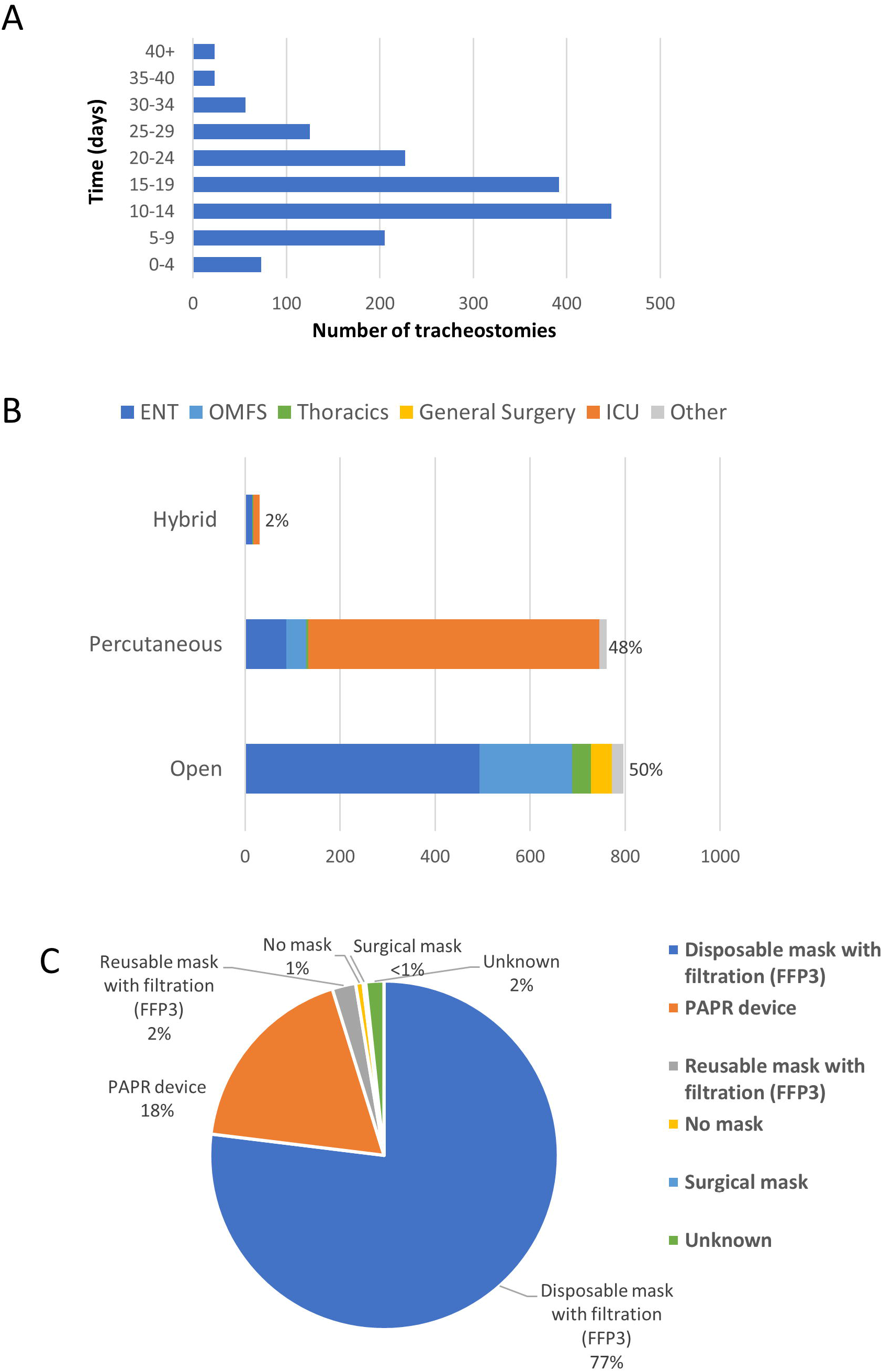
Length of time from intubation to tracheostomy (days) **Figure 2b** Number of tracheostomies divided by method and specialist performing the procedure. **Figure 2c**. Type of respirator used during the tracheostomy.

### Tracheostomy procedure

“Anticipated prolonged wean” was the most cited indication for tracheostomy in 1473 (92%). An open method of tracheostomy was used in 797 (50%) procedures, percutaneous method in 771 (48%), and a hybrid method using a combination of open and percutaneous techniques in 31 (2%) (Figure 2b). Bronchoscopy was used as an aid in 574 (78%) percutaneous techniques. Patient factors likely to make the tracheostomy more challenging were reported in 327 (41%) open and 109 (14%) percutaneous tracheostomies. Of these, neck obesity was the most frequently reported, occurring in 242 (30%) of all open tracheostomies.

Operators used either an FFP3 mask or Powered Air Purifying Respirator (PAPR) in 1563 (99%) of cases (Figure 2c). Other PPE included double gloves in 1460 (91%), a surgical gown in 1511 (94%), and a face visor was used in 1261 (96%) of cases in addition to an FFP3 mask.

### Complications

Intraoperative complications were reported in 147 (9%) procedures with oxygen desaturation below 80% being the most common (n=67), followed by intra-operative bleeding (n=29). Post-operative complications occurred in 356 (22%) cases; bleeding was reported in 119 (7%) patients, more frequently following open procedures (p<0.001). A leak around the tracheostomy cuff necessitating a change in tube was reported in 75 (5%) cases, 48 of these were open tracheostomies.

### Outcomes

In this cohort, one patient died during the tracheostomy procedure, 276 (17%) patients died before weaning from mechanical ventilation and a further nine (1%) patients died between successful weaning and hospital discharge (Table 3). 249 (89%) deaths were COVID-19 related and nine patients died of tracheostomy-related complications.

**Table 3.**
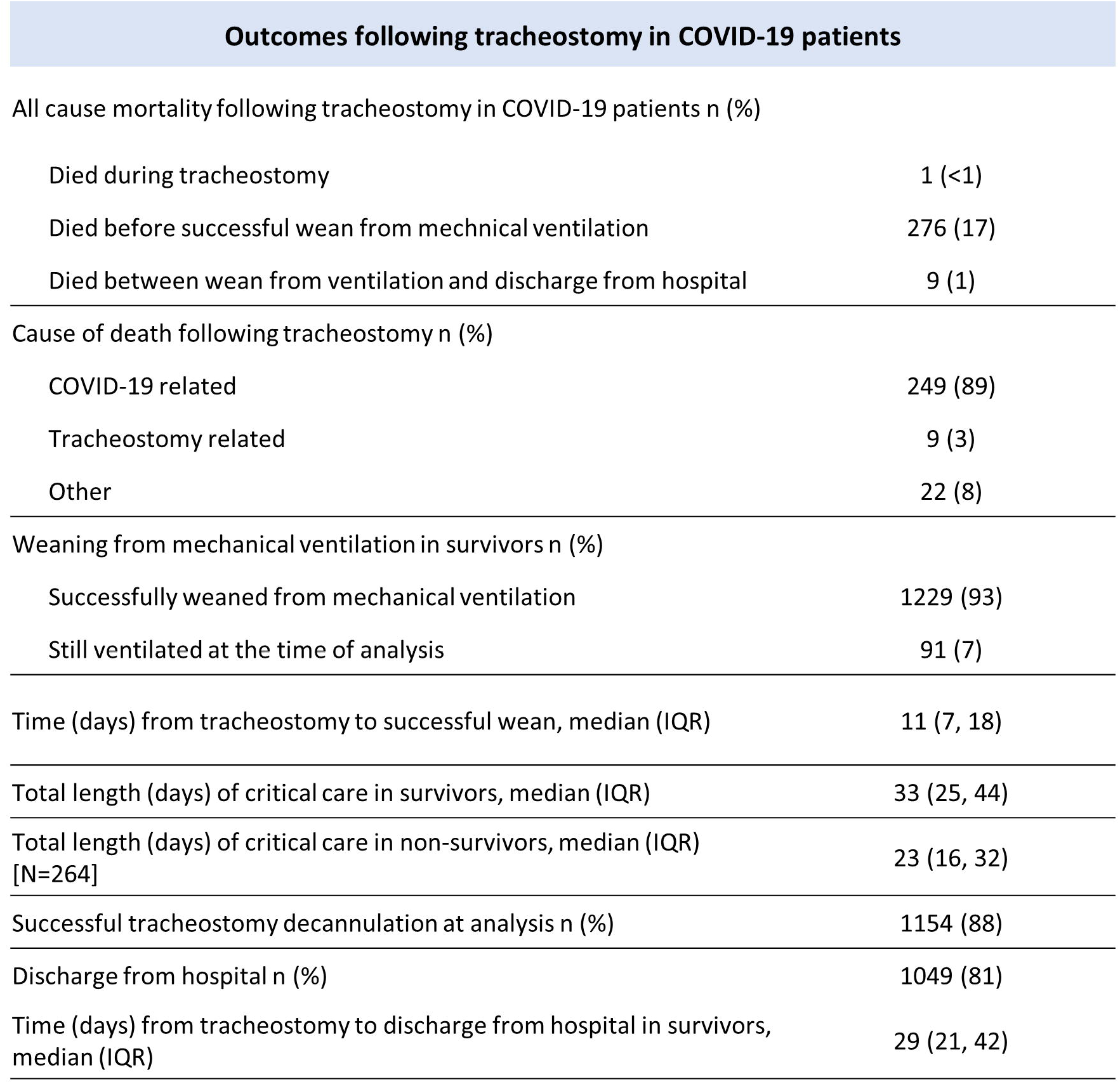
Outcomes of mechanically ventilated patients with COVID-19 undergoing tracheostomy.

Characteristics of survivors and non-survivors are shown in Supplementary Table 2. A multivariable logistic regression model was fitted for mortality and backward elimination applied. Age, days of mechanical ventilation pre-procedure, inspired oxygen concentration and PEEP setting at the time of tracheostomy, use of inotropic support (all p<0.001), upward trending CRP (P=0.003), pyrexia (p=0.003) and use of anticoagulation (p=0.002) are independently associated with mortality (Figure 3a).

**Figure 3a.**
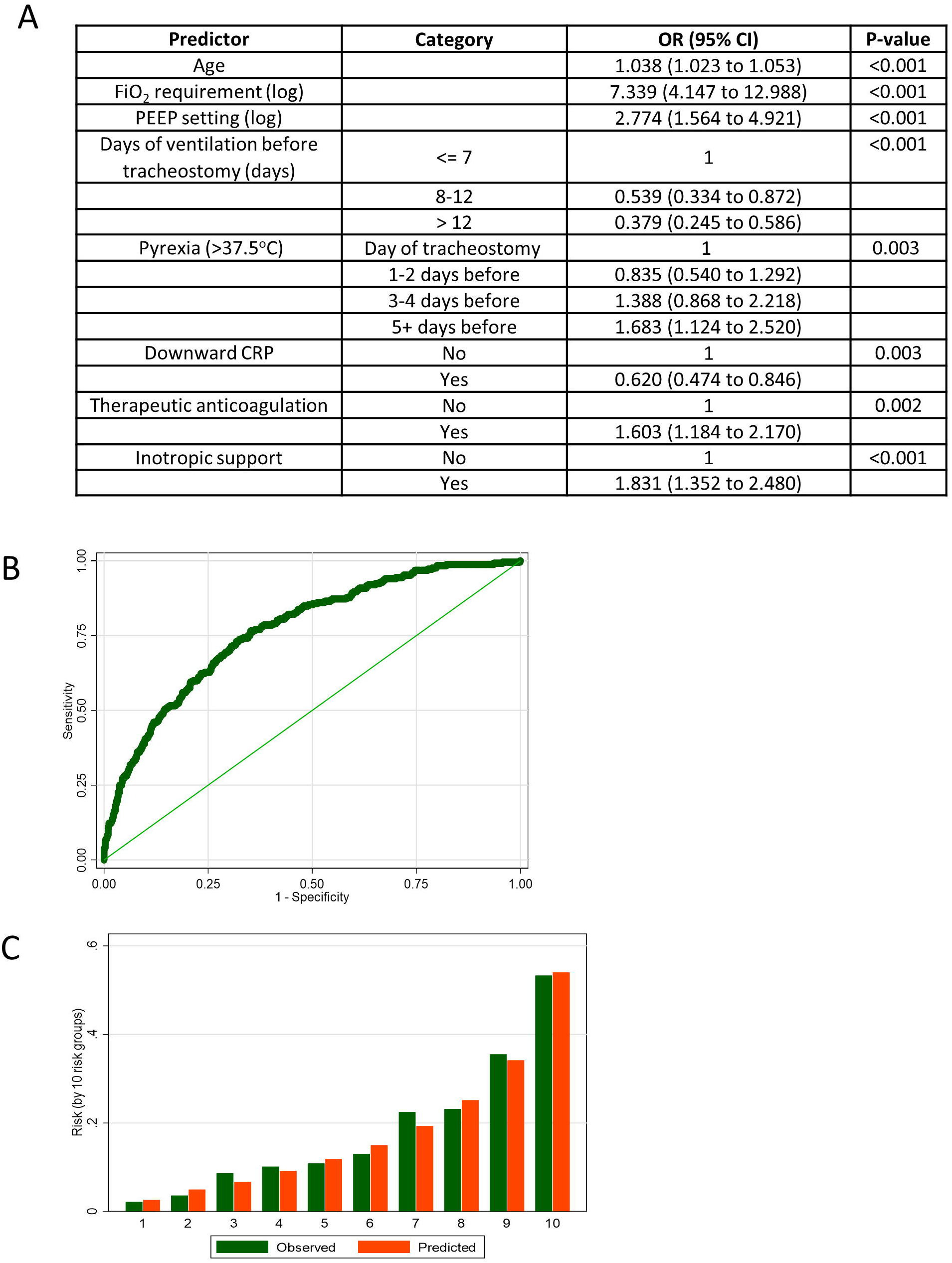
Odds ratios from multivariable logistic regression model for mortality (n=1379). Eight predictive clinical variables were identified. **Figure 3b** ROC curve for multivariable prediction model. The optimal cut-point for sensitivity and specificity, found using Youden’s index, is based on a predicted value of 15.8%. This provides a sensitivity of 77% and specificity of 65%. The corresponding PPV and NPV are 33% and 93% respectively. **Figure 3c** Observed and predicted risk for 10 quantile groups. The groups (1-10) were obtained by splitting patients by their predicted risk, i.e. group 1 comprise the 10% of patients with the lowest predicted risk.

A model was developed to predict mortality. Internal validation using 10-fold cross-validation produced an average ROC area of 0.76 (95% CI: 0.73 to 0.79), suggesting good discrimination (Figure 3b). The Hosmer-Lemeshow test results in each cross-validation fold suggest no problems with calibration. A comparison of observed and predicted outcomes suggests good agreement (Figure 3c) with a sensitivity of 77% and specificity of 65%. The corresponding positive predictive value and negative predictive value are 33% and 93% respectively.

Of the survivors, 1229 (93%) had been successfully weaned at the time of analysis with 1154 (88%) having undergone successful tube decannulation and 1049 (81%) discharged from hospital (Table 3). Median time from tracheostomy to discharge to hospital in survivors was 29 days (IQR 21, 42).

Characteristics of patients according to length of time to successful wean from ventilation are shown in Supplementary Table 3. A multivariable logistic regression model for time to wean success was fitted and backward elimination applied. These results suggest that advanced age, male sex, higher PEEP setting, higher inspired oxygen requirement, use of anticoagulation (all p<0.001) and non-invasive ventilation before tracheostomy (p=0.003) were all independently associated with prolonged periods of ventilation following tracheostomy (Table 4). An association was found between insertion technique and time to successful wean, although the difference was small (median 12 days for percutaneous versus 11 days for open method). No association was found between time from intubation to tracheostomy and time from tracheostomy to successful ventilatory wean (p=0.92).

**Table 4.** Regression coefficients from multiple regression model for time to wean success (n=1039)

| Predictor | Category | Coefficient (95% CI) | P-value |
| --- | --- | --- | --- |
| Age |  | 0.010 (0.007 to 0.014) | <0.001 |
| Sex | Male | 0 | 0.025 |
|  | Female | -0.101 (-0.189 to -0.012) |  |
| PEEP setting (log) |  | 0.288 (0.147 to 0.428) | <0.001 |
| FiO2 requirement (log) |  | 0.396 (0.236 to 0.556) | <0.001 |
| Method of tracheostomy | Open | 0 | 0.010 |
|  | Percutaneous | 0.110 (0.027 to 0.193) |  |
| Anticoagulant | No | 0 | <0.001 |
|  | Yes | 0.157 (0.074 to 0.240) |  |
| Use of NIV | No | 0 | 0.003 |
|  | Yes | 0.124 (0.041 to 0.207) |  |
| Intercept |  | -0.246 (-0.868 to 0.376) | 0.438 |

### SARS-CoV-2 infection in operators

The question “Did any of the operators test positive for COVID-19 within two weeks of the procedure?”, was answered in 97% (1558/1605) of cases. Six instances were reported across four hospitals, four after percutaneous tracheostomy and two after open tracheostomy. Five of the cases were performed in intensive care and four within a negative pressure environment. Personal protective equipment used in these cases included an FFP3 mask in four cases, a fluid-resistant hood with face visor in one, and a PAPR device in one.

## Discussion

This study involved 126 UK hospitals reporting on 1605 individual tracheostomies. Given the Intensive Care National Audit reports on 7792 patients requiring advanced respiratory support across 265 hospitals in England, Wales and Northern Island, our cohort is representative, and the results are likely generalizable to the UK.

At the time of censoring, all-cause mortality following tracheostomy in our cohort was 18%. This number is likely to rise as 91 patients were still mechanically ventilated and a further 171 had been weaned but were still in hospital. Prospective multicentre studies of general (non-COVID-19) intensive care populations patients report mortality rates of approximately 30% in the first 30 days following tracheostomy [24, 25]. Direct comparisons to this cohort of COVID-19 patients cannot however be drawn as demographics, comorbidities and underlying pathologies will differ considerably and timing of tracheostomy is usually performed earlier than the median 15 days following intubation reported in this study of COVID-19 patients. National data reported in the ICNARC registry, indicates that the ICU mortality rate in mechanically ventilated COVID-19 patients was 47.8%, however median duration of critical care stay in non-survivors was 10 days (IQR 6,17) [7]. Thus COVID-19 patients undergoing tracheostomy constitute a preselected population who have survived the acute phase and, in general, would have cardiorespiratory stability and are no longer requiring high-level ventilatory support and high inspired oxygen concentrations. Nonetheless, our data shows that tracheostomy in the setting of SARS-CoV-2 infection is not a futile intervention as previously claimed by some expert opinion at the start of the pandemic [26, 27].

Whether the timing of tracheostomy does influence COVID-19 patient outcomes is unclear. Early tracheostomy may benefit certain patient groups [28, 29], but meta-analyses have failed to show benefit in a general population of critically ill adults [30, 31]. In randomized trials, approximately half of patients randomized to late tracheostomy eventually did not undergo the procedure, mainly due to successful extubation [24, 31]. In our cohort, early tracheostomy was independently associated with higher mortality. Moreover, no association was demonstrated between early tracheostomy and shortened time to successful weaning from ventilation. Clearly, cause and effect cannot be directly inferred from this data and only prospective randomized studies could address this important question.

We found no association between method of tracheostomy and likelihood of successful wean from ventilation, mortality, or discharge from hospital. As in non-COVID-19 series, bleeding was more frequent using the open method, although the overall rate of reported bleeding was low. The percutaneous method has several advantages centered around the ability to perform the procedure at the bedside. In contrast, the open method enables safe procedure in those with difficult neck anatomy and enables the surgical workforce to relieve the task from intensive care staff during periods when a critical care department is working at full capacity. The decision over which method to employ should be locally led and depends upon expertise available and close inter-disciplinary working.

The low rates of reported SARS-COV-2 infection among operators who likely continued to work in other high-risk areas and performed other aerosol generating procedures is encouraging. Whilst asymptomatic cases may have been missed and recall bias may have occurred, the low rates of infection suggest that, with appropriate PPE, the procedure does not pose a high risk of infection with SARS-COV-2 to operators. Our findings are consistent with other series [32, 33].

Infectivity and viral load is believed to peak around the time of symptom onset and then decline over the following 3-4 days [34, 35]. Considering the median time from symptom presentation to hospitalization is four days and that tracheostomy is not usually considered until at least seven days after intubation, the risk of infectivity is predicted to be low even if the procedure is performed between the first and second week of ventilation [4, 36]. Our results therefore do not support guidance suggesting tracheostomy should be delayed until 14-21 days after intubation to reduce the potential for infection amongst operators [14, 16]. Similarly, our findings and data showing a positive COVID-19 test does not correlate with risk of infectivity later in the disease process, suggest tracheostomy should not be delayed to achieve a negative COVID-19 test. Delaying in these circumstances defers the potential benefits of tracheostomy and increases the risk of complications relating to prolonged endotracheal intubation without any clear benefit to the patient or operators involved in the procedure.

## Data Availability

The authors confirm that the data supporting the findings of this study are available within the article and its supplementary materials

## Authors contributions

NH contributed to literature search, figures, study design, data collection, analysis, interpretation, and writing. AS contributed to study design, data analysis and interpretation and writing. GA contributed to data analysis, interpretation, and writing. TJ contributed to study design, data interpretation and writing. MS contributed to data analysis & interpretation and writing. All other authors contributed equally under the principles of corporate authorship.

## Ethics committee approval

COVIDTrach has been approved by the UK Research and Ethics Council and the Health Research Authority as a prospective cohort study.

## Data collection

Study data were collected and managed using REDCap electronic data capture tools hosted at University College London.[37, 38] REDCap (Research Electronic Data Capture) is a secure, web-based application designed to support data capture for research studies, providing: 1) an intuitive interface for validated data entry; 2) audit trails for tracking data manipulation and export procedures; 3) automated export procedures for seamless data downloads to common statistical packages; and 4) procedures for data integration and interoperability with external sources.

---

**Supplementary Table 1.** Data completeness for each variable examined with total number of missing answers.

| Variable | % answered | Total number expected | Total number completed | Number missing |
| --- | --- | --- | --- | --- |
| Age | 100 | 1605 | 1605 | 0 |
| Gender | 100 | 1605 | 1604 | 1 |
| COVID status | 100 | 1605 | 1599 | 6 |
| Method of tracheostomy | 100 | 1605 | 1598 | 7 |
| Mortality | 100 | 1605 | 1601 | 4 |
| Wean from ventilator | 100 | 1326 | 1320 | 6 |
| CRP before tracheostomy | 99 | 1605 | 1583 | 22 |
| Location of tracheostomy | 99 | 1605 | 1594 | 11 |
| Surgical specialty performing operation | 99 | 1605 | 1588 | 17 |
| Intraoperative complications | 99 | 1605 | 1581 | 24 |
| Discharge from hospital within 28 days | 99 | 1326 | 1315 | 11 |
| Intubation to tracheostomy (days) | 98 | 1605 | 1565 | 40 |
| Inotropic support | 98 | 1605 | 1565 | 40 |
| Indication for tracheostomy | 98 | 1605 | 1569 | 36 |
| Neck factors affecting tracheostomy | 98 | 1605 | 1567 | 38 |
| Use of PPE - mask use | 98 | 1605 | 1578 | 27 |
| FiO2 on day of tracheostomy | 97 | 1605 | 1549 | 56 |
| Cause of death | 97 | 299 | 291 | 8 |
| COVID-19 amongst operators | 97 | 1605 | 1558 | 47 |
| PEEP on day of tracheostomy | 96 | 1605 | 1533 | 72 |
| Guidance method for perc tracheostomy | 96 | 771 | 736 | 35 |
| Co-morbidities | 95 | 1605 | 1521 | 84 |
| Admission to intubation (days) | 95 | 1605 | 1518 | 87 |
| Use of NIV | 95 | 1605 | 1525 | 80 |
| Postoperative complications before wean | 95 | 1605 | 1530 | 75 |
| Total length of critical care in non-survivors | 95 | 279 | 264 | 15 |
| Type of tracheostomy tube | 94 | 1605 | 1509 | 96 |
| Temperature before tracheostomy | 92 | 1605 | 1481 | 124 |
| P02 on day of tracheostomy | 92 | 1605 | 1480 | 125 |
| Outcome of last test | 91 | 1605 | 1455 | 150 |
| BMI | 90 | 1605 | 1437 | 168 |
| Number of tests | 90 | 1605 | 1441 | 164 |
| Time from test to tracheostomy (days) | 90 | 1605 | 1451 | 154 |
| Tracheostomy decannulation | 90 | 1326 | 1195 | 131 |
| Time from tracheostomy to wean | 88 | 1326 | 1164 | 162 |
| Postoperative complications after wean | 86 | 1326 | 1142 | 184 |
| Total length of critical care in survivors | 85 | 1326 | 1121 | 205 |

**Supplementary Table 2.** Baseline and demographic characteristics by mortality. Values are either N (%) or mean (SD)

|  | Survived (n=1319) | Died (n=286) |
| --- | --- | --- |
| Age |  |  |
| <30 | 22 (88.0%) | 3 (12.0%) |
| 30-49 | 293 (89.6%) | 34 (10.4%) |
| 50-69 | 854 (82.9%) | 176 (17.1%) |
| 70+ | 150 (67.3%) | 73 (32.7%) |
| Sex |  |  |
| Male | 909 (81.0%) | 213 (19.0%) |
| Female | 409 (84.9%) | 73 (15.1%) |
| Missing | 1 | 0 |
| Body Mass Index |  |  |
| <18.5 | 12 (92.3%) | 1 (7.7%) |
| 18.5-25 | 286 (80.6%) | 69 (19.4%) |
| 25-30 | 395 (82.6%) | 83 (17.4%) |
| 30-40 | 391 (83.5%) | 77 (16.5%) |
| 40+ | 109 (89.3%) | 13 (10.7%) |
| Missing | 126 | 43 |
| Number of Comorbidities |  |  |
| None | 390 (82.3%) | 84 (17.7%) |
| One | 458 (81.3%) | 105 (18.7%) |
| Two or more | 471 (82.9%) | 97 (17.1%) |
| CRP | 112.0 (84.6) | 151.4 (98.4) |
| Missing | 19 | 3 |
| Downward CRP |  |  |
| No | 388 (77.4%) | 113 (22.6%) |
| Yes | 904 (84.5%) | 166 (15.5%) |
| Missing | 27 | 7 |
| Anticoagulant |  |  |
| No | 700 (84.7%) | 126 (15.3%) |
| Yes | 546 (78.2%) | 152 (21.8%) |
| Missing | 73 | 8 |
| Inotropic support |  |  |
| No | 833 (86.9%) | 126 (13.1%) |
| Yes | 456 (75.2%) | 150 (24.8%) |
| Missing | 30 | 10 |
| Use of NIV |  |  |
| No | 721 (82.5%) | 153 (17.5%) |
| Yes | 535 (82.2%) | 116 (17.8%) |
| Missing | 63 | 17 |
| Days of ventilation before tracheostomy |  |  |
| <= 7 | 99 (63.1%) | 58 (36.9%) |
| 8 to 12 | 293 (80.1%) | 73 (19.9%) |
| > 12 | 895 (86.0%) | 146 (14.0%) |
| Missing | 32 | 9 |
| FiO2 requirement | 38.5 (11.1) | 46.7 (13.7) |
| Missing | 47 | 9 |
| PO2 requirement | 10.9 (7.1) | 11.2 (10.6) |
| Missing | 112 | 13 |
| PEEP setting | 7.9 (2.4) | 8.9 (2.1) |
| Missing | 59 | 13 |
| Method of tracheostomy |  |  |
| Open | 681 (85.6%) | 115 (14.4%) |
| Percutaneous | 614 (79.6%) | 157 (20.4%) |
| Missing / Hybrid | 24 | 14 |

**Supplementary Table 3.** Days to Wean Success. Values are Spearman correlation coefficient or median (inter-quartile range). P-values are from either spearman correlation or Kruskal-Wallis test.

| Varibale | N | Days (n=1165) | P |
| --- | --- | --- | --- |
| Age |  |  | <0.001 |
| <30 | 18 | 8.5 (7 to 12) |  |
| 30-49 | 260 | 10 (6 to 15) |  |
| 50-69 | 753 | 12 (7 to 18) |  |
| 70+ | 134 | 13 (8 to 23) |  |
| Sex |  |  | <0.001 |
| Male | 798 | 12 (7 to 19) |  |
| Female | 366 | 9.5 (6 to 16) |  |
| Missing | 1 |  |  |
| Body Mass Index |  |  | 0.023 |
| <18.5 | 12 | 11 (6 to 22) |  |
| 18.5-25 | 260 | 13 (7 to 22) |  |
| 25-30 | 356 | 11 (7 to 17.5) |  |
| 30-40 | 349 | 11 (7 to 17) |  |
| 40+ | 97 | 10 (7 to 15) |  |
| Missing | 91 |  |  |
| Number of Comorbidities |  |  | 0.521 |
| None | 351 | 11 (6 to 18) |  |
| One | 394 | 12 (7 to 18) |  |
| Two or more | 420 | 11 (7 to 17) |  |
| CRP | 1152 | 0.11 | <0.001 |
| Missing | 13 |  |  |
| Downward CRP |  |  | 0.574 |
| No | 338 | 12 (7 to 19) |  |
| Yes | 807 | 11 (7 to 18) |  |
| Missing | 20 |  |  |
| Anticoagulant |  |  | <0.001 |
| No | 628 | 10 (6 to 16) |  |
| Yes | 492 | 12 (8 to 21) |  |
| Missing | 45 |  |  |
| Inotropic support |  |  | 0.002 |
| No | 757 | 11 (6 to 18) |  |
| Yes | 389 | 12 (8 to 19) |  |
| Missing | 19 |  |  |
| Use of NIV |  |  | <0.001 |
| No | 644 | 10 (6 to 17) |  |
| Yes | 486 | 12 (8 to 20) |  |
| Missing | 35 |  |  |
| Days of ventilation before tracheostomy |  |  | 0.107 |
| <= 7 | 83 | 12 (7 to 17) |  |
| 08-Dec | 265 | 11 (7 to 18) |  |
| > 12 | 810 | 11 (7 to 18) |  |
| Missing | 7 |  |  |
| FiO2 requirement | 1145 | 0.21 | <0.001 |
| Missing | 20 |  |  |
| PO2 requirement | 1096 | -0.07 | 0.022 |
| Missing | 69 |  |  |
| PEEP setting | 1136 | 0.17 | <0.001 |
| Missing | 29 |  |  |
| Method of tracheostomy |  |  | 0.096 |
| Open | 586 | 11 (6 to 18) |  |
| Percutaneous | 557 | 12 (7 to 18) |  |
| Missing / Hybrid | 22 |  |  |

## References

1. Worldometer. COVID-19 Coronavirus Pandemic. 2020 [cited 2020 10.09.20]; Available from: https://www.worldometers.info/coronavirus/.

2. Wu, Z. and J.M. McGoogan, Characteristics of and Important Lessons From the Coronavirus Disease 2019 (COVID-19) Outbreak in China: Summary of a Report of 72314 Cases From the Chinese Center for Disease Control and Prevention. JAMA, 2020. 323(13): p. 1239–1242.

3. Grasselli, G., A. Pesenti, and M. Cecconi, Critical Care Utilization for the COVID-19 Outbreak in Lombardy, Italy: Early Experience and Forecast During an Emergency Response. JAMA, 2020. 323(16): p. 1545–1546.

4. Cummings, M.J., et al., Epidemiology, clinical course, and outcomes of critically ill adults with COVID-19 in New York City: a prospective cohort study. Lancet, 2020. 395(10239): p. 1763–1770.

5. Grasselli, G., et al., Baseline Characteristics and Outcomes of 1591 Patients Infected With SARS-CoV-2 Admitted to ICUs of the Lombardy Region, Italy. JAMA, 2020. 323(16): p. 1574–1581.

6. Suleyman, G., et al., Clinical Characteristics and Morbidity Associated With Coronavirus Disease 2019 in a Series of Patients in Metropolitan Detroit. JAMA Netw Open, 2020. 3(6): p. e2012270.

7. Centre, I.C.N.A.R. ICNARC report on COVID-19 in critical care in Egnland, Wales and Northern Ireland 25 September 2020. 2020 [cited 2020 28.09.20]; Available from: https://www.icnarc.org/Our-Audit/Audits/Cmp/Reports.

8. Durbin, C.G., Jr., M.P. Perkins, and L.K. Moores, Should tracheostomy be performed as early as 72 hours in patients requiring prolonged mechanical ventilation? Respir Care, 2010. 55(1): p. 76–87.

9. Groves, D.S. and C.G. Durbin, Jr., Tracheostomy in the critically ill: indications, timing and techniques. Curr Opin Crit Care, 2007. 13(1): p. 90–7.

10. Jaeger, J.M., K.A. Littlewood, and C.G. Durbin, Jr., The role of tracheostomy in weaning from mechanical ventilation. Respir Care, 2002. 47(4): p. 469-80; discussion 481-2.

11. Pierson, D.J., Tracheostomy and weaning. Respir Care, 2005. 50(4): p. 526–33.

12. Miles, B.A., et al., Tracheostomy during SARS-CoV-2 pandemic: Recommendations from the New York Head and Neck Society. Head Neck, 2020. 42(6): p. 1282–1290.

13. ENT.UK. COVID-19 Tracheostomy Guidance. 2020 [cited 2020 11.04.20]; Available from: https://www.entuk.org/covid-19-tracheostomy-guidance-t-jacob-et-al.

14. Sommer, D.D., et al., Recommendations from the CSO-HNS taskforce on performance of tracheotomy during the COVID-19 pandemic. J Otolaryngol Head Neck Surg, 2020. 49(1): p. 23.

15. Association, B.L. Tracheostomy Guideline COVID-19. 2020 [cited 2020 05.05.20]; Available from: https://www.britishlaryngological.org/news/tracheostomy-guideline-covid-19.

16. Michetti, C.P., et al., Performing tracheostomy during the Covid-19 pandemic: guidance and recommendations from the Critical Care and Acute Care Surgery Committees of the American Association for the Surgery of Trauma. Trauma Surg Acute Care Open, 2020. 5(1): p. e000482.

17. Heyd, C.P., et al., Tracheostomy protocols during COVID-19 pandemic. Head Neck, 2020. 42(6): p. 1297–1302.

18. Zou, L., et al., SARS-CoV-2 Viral Load in Upper Respiratory Specimens of Infected Patients. N Engl J Med, 2020. 382(12): p. 1177–1179.

19. McGrath BA, B.M., Warrillow SJ, Pandian V, Arora A, Cameron TS, Anon JM, Martinez GH, Truog RD, Block SD, Lui GCY, McDonald C, Rassekh CH, Atkins J, Qiang L, Vergez S, Dulguerov P, Zenk J, Antonelli M, Pelosi P, Walsh BK, Ward E, Shang Y, Gasparini S, Donati A, Singer M, Openshaw PJM, Tolley N, Markel H, Feller-Kopman DJ. Tracheostomy in the COVID-19 era: global and multidisciplinary guidance. Lancet Resp Med. 2020; doi.org/10.1016/S2213-2600(20)30230230-7.

20. Chao, T.N., et al., Outcomes after Tracheostomy in COVID-19 Patients. Ann Surg, 2020.

21. Martin-Villares, C., et al., Outcome of 1890 tracheostomies for critical COVID-19 patients: a national cohort study in Spain. Eur Arch Otorhinolaryngol, 2020.

22. Angel, L., et al., Novel Percutaneous Tracheostomy for Critically Ill Patients With COVID-19. Ann Thorac Surg, 2020. 110(3): p. 1006–1011.

23. collaborative, C.O., COVIDTrach; the outcomes of mechanically ventilated COVID-19 patients undergoing tracheostomy in the UK: Interim Report. Br J Surg, 2020.

24. Young, D., et al., Effect of early vs late tracheostomy placement on survival in patients receiving mechanical ventilation: the TracMan randomized trial. JAMA, 2013. 309(20): p. 2121–9.

25. Terragni, P.P., et al., Early vs late tracheotomy for prevention of pneumonia in mechanically ventilated adult ICU patients: a randomized controlled trial. JAMA, 2010. 303(15): p. 1483–9.

26. Vukkadala, N., et al., COVID-19 and the Otolaryngologist: Preliminary Evidence-Based Review. Laryngoscope, 2020.

27. Lavinsky, J., et al., An update on COVID-19 for the otorhinolaryngologist - a Brazilian Association of Otolaryngology and Cervicofacial Surgery (ABORL-CCF) Position Statement. Braz J Otorhinolaryngol, 2020. 86(3): p. 273–280.

28. Wang, R., et al., The impact of tracheotomy timing in critically ill patients undergoing mechanical ventilation: A meta-analysis of randomized controlled clinical trials with trial sequential analysis. Heart Lung, 2019. 48(1): p. 46–54.

29. Elkbuli, A., et al., Early versus Late Tracheostomy: Is There an Outcome Difference? Am Surg, 2019. 85(4): p. 370–375.

30. Huang, H., et al., Timing of tracheostomy in critically ill patients: a meta-analysis. PLoS One, 2014. 9(3): p. e92981.

31. Meng, L., et al., Early vs late tracheostomy in critically ill patients: a systematic review and meta-analysis. Clin Respir J, 2016. 10(6): p. 684–692.

32. Riestra-Ayora, J., et al., Safety and Prognosis in Percutaneous vs Surgical Tracheostomy in 27 Patients With COVID-19. Otolaryngol Head Neck Surg, 2020. 163(3): p. 462–464.

33. Williamson, A., et al., Early percutaneous tracheostomy for patients with COVID-19. Anaesthesia, 2020.

34. Wolfel, R., et al., Virological assessment of hospitalized patients with COVID-2019. Nature, 2020. 581(7809): p. 465–469.

35. McGrath, B.A., et al., Tracheostomy in the COVID-19 era: global and multidisciplinary guidance. Lancet Respir Med, 2020. 8(7): p. 717–725.

36. Zhou, F., et al., Clinical course and risk factors for mortality of adult inpatients with COVID-19 in Wuhan, China: a retrospective cohort study. Lancet, 2020. 395(10229): p. 1054–1062.

37. Harris, P.A., et al., The REDCap consortium: Building an international community of software platform partners. J Biomed Inform, 2019. 95: p. 103208.

38. Harris, P.A., et al., Research electronic data capture (REDCap)--a metadata-driven methodology and workflow process for providing translational research informatics support. J Biomed Inform, 2009. 42(2): p. 377–81.

